# Analysis of the association between *rs111200466* variants and cervical Cancer susceptibility in Sudanese women

**DOI:** 10.1101/2021.07.05.21250955

**Authors:** Safaa Othman Andarawi, Sahar Bakheit

**Author notes:** 00249912681836.

## Abstract

Worldwide there are more than 273,000 deaths from cervical cancer each year and it accounts for 9% of female cancer deaths. The high-risk variants of human papillomavirus such as HPV 16 and 18 are major etiological agents of cervical cancer. The persistency of infection with HPV may due to inappropriate immune response due to SNPs interfering with the function of particular part of the immune system. Toll-like receptors (TLRs) play an important role in the signaling of many pathogen-related molecules and endogenous proteins associated with immune activation. The –196 to –174del polymorphism affects the TLR2 gene and alters its promoter activity.

**Aim:** In this study we investigated the presence and the influence of TLR2 –196 to – 174del polymorphism on the risk of cervical cancer development in Sudanese. The study was performed on 42 patients with cervical cancer and 27 healthy controls. Genotyping of –196 to –174del polymorphism of TLR2 was investigated using allele-specific polymerase chain reaction method in all subjects. We documented the presence of–196 to –174del polymorphism of TLR2 in Sudanese women with allele frequency 37% of control and 34% of patient sample although there was no observed association between this SNP and the risk of cervical cancer (p value. 394).Further studies are needed in a large and ethnically diverse population to determine the impact of the TLR2 polymorphism in the susceptibility of cervical cancer.

## Introduction and literature review

### Epidemiology of cervical cancer

Cervical carcinoma is the primary cause of cancer-related deaths amongst women in the majority of developing countries and the second most prominent cancer among women worldwide. Every year approximately 371000 invasive cervical carcinoma diagnosis worldwide that accounts for 10% of all neoplastic disease in women. In frequency, it is the seventh cancer site overall and third among women, after breast and colorectal cancer. In developing countries, until the early 1990 cervical carcinoma was the numerous neoplastic diseases, after which breast cancer became the major neoplastic disease.(1).

### Human papillomavirus and cervical cancer

There are two main types of cervical cancer - squamous cell cancer called carcinoma (the most common) and adenocarcinoma, although they could be mixed.

The fact that the exclusive cause of cervical cancer is HPV is the results of large and constant finding of researches carried out along several years as early as 1980. This studies has a clear result indicating a strong and specific role of the viral infection in all settings where investigations have taken place. The result of amplification techniques used in case-control studies, case-series and prevalence surveys approve that, in satisfactory specimens of cervical cancer, HPV DNA can be detected in 90 to 100% of the cases. (2).

Many several researches suggested that the majority of the cases is developed after persistent infection with the high risk types of HPV (3)The viruses are species-specific, and also exquisitely tissue-tropic, with a preference to infect either cutaneous or internal squamous mucosal surfaces. Liaw and his collogues in 2001 documented that” Papillomaviruses are not classified by serotype, but by genotype. (4)

The low risk HPV associated with benign anogenital warts or condylomata include (HPV 6,11 and their relatives). While the high risk types are (HPV 16, 18, 31, 33, 35 and 45).these high risk types are associated with anogenital cancers and the intra epithelial neoplasial lesion particularly of the cervix. The result of a research carried by Mockenhaupt in 2006 concluded that “almost 100% of cervical cancers, the second commonest cancer in women worldwide, contain the HPV DNA sequences from a high-risk oncogenic genital HPV. The major players are HPV16, found in 50–70% of cases, and HPV18 detected in 7–20% of cases”(5).

### Cervical cancer in Sudan

cervical cancer in Sudan represent 9.8% of female genital cancer which is not reflect the real percentage as only 10%of cases reach the hospital. incidence of cervical cancer in Sudan is more than21.0,..only 10%of cases were screened in the past 5 years, annual mortality is more than the mortality of pregnancy and child berth (6).

### HPV viral persistence

Repeated sampling of women being followed for viral persistence and cervical abnormalities has shown that the median duration of the infections is around 8 months for high risk HPV types as compared to 4.8 months for the low-risk HPV types. In a study in of high-risk population in Brazil, the mean duration of HPV detection was 13.5 months for high-risk HPV types and 8.2 months for the non-oncogenic types. HPV 16 tended to persist longer than the average for high-risk types other than HPV 16 (7).

### The biology of HPV infection

Papillomaviruses are double-stranded DNA viruses, which replicate exclusively in stratified squamous epithelia, using the differentiation of the epithelium to regulate their replication. Virions targets the basal layer stem cells of the epithelia. After invasion of the layers through mirobrasions the virus replicate episomally using the cellular DNA replication machinery and Ei,E2 virus nonstructural proteins. As a result of viral nonstructural proteins E 6 and E 7 expression, the cell cycle arrest and differentiation is delayed. following this delay the virus can replicate more using host replication machinery in the suprabasal epithelial cells causing the thickening of the skin the typical feature of some papillomavirus infection. When the replicating epithelial cells differentiate to non-replicating mature keratinocytes in the nucleus the late L1 and L 2 assembled followed by the release of mature virions from the epithelium.

### Innate immunity and viral infection

Pattern recognition receptors is the main player in the detection of viral particles or DNA, RNA by the innate immune cells. After recognition of the pathogen PRRs is known to induce INFs and other proliferative cytokines in the infected cells and other immune cells. There are varieties of these receptor the will know family are members of Toll like receptors (8).

“Mammalian TLRs play a key role in host defense during pathogen infection byregulating and linking the innate and adaptive immune responses” (9) four of the known mammalian TLRs have been implicated in nucleic acid sensing: TLR3, TLR7, TLR8, and TLR9.

### Functional consequences of TLR activation during viral infection

Following viral recognition by TLR in specific cells like dendritic cells, subsequently series of signal transduction will contribute to a wide range of antiviral response this include expression of proinflammatory cytokines, activation of antigen presenting cells such as dendritic cells and macrophages, NK cell activation, and induction of adaptive immunity..

#### Toll like receptor 2

TLR has been reported in many study as immune system activator after virus recognition. The haemaglutinin (HA) protein of wild type, but not vaccine strains of measles virus activates murine and human cells via TLR2, leading to induction of proinflammatory cytokines, such as IL-6 in human monocytic cells, and up regulation of surface expression of CD150, a receptor for measles virus (10).Activation of TLR2-dependent signaling by wild type measles virus is likely contribute to both immune activation and viral spread and pathogenicity.. Human cytomegalovirus (HCMV) has also been shown to mediate cellular effects via TLR2. In cells lacking TLR2 or CD14 (a co-receptor for TLR2 and TLR4), UV-inactivated HCMV virions could no longer activate NFºB nor induce IL-6 and IL-8. Compton suggested that “the loss of TLR2-activating ability may be indicate as an attenuation marker” (11)

In contrast to what have been suggested by many researches that TLR2 activation is essential for viral immune reaction Rajagopal and his collogue concluded that although TLR2 signaling is essential for the production of: TNFα, IL-1β, IL-6, IL-12, CCL7,against HSV. However, TLR2-/-mice display significantly reduced mortality and ceased neuroinflammation in response to brain infection with HSV(12)

### Genetic polymorphisms in Toll-like receptors

By reviewing the literature its clear that TLR mutations have been intensively studied and many of these studies suggested that present of these SNPs may configure considerable change toward progression of inflammatory and infectious diseases. The three most common TLR2 SNPs are −196 to −174 del polymorphism, Arg677Trp and Arg753Gln.

Kang and his colloquies studied Arg 677Trp mutation and concluded that, thisSNP has a crucial effect in the intracellular domain function of TLR2.the effect led to diminishing the production of cytokines mainly IL2.they suggested that this defect is due to failure to interact with MyD88.Another finding of their study is that this polymorphism associated with lepromatous leprosy in a Korean population (13)

In a similar study this polymorphism found to be associated with repeated bacterial infections in Turkish children(14).In Addison to these association, (15)has recently reported that The −196 to −174 del polymorphism in TLR 2 gene located on chromosome 4, change the promoter activity of TLR 2. He concluded that “significant association between the del allele of TLR 2 and the risk of developing cervical cancer “. -196 to -174del has intensively studied in the context of inflammation and infectious diseases. in 2010 Alisa reveals that there is association between the TLR2 -196 to -174 del and asymptomatic bancroftianfilariasis (p-value 0.0011))(16).In 2010yang evaluate the effect -196 to -174 del in TB infection and its severity, and they concluded that it is associated with a systemic TB symptoms (17)

## Material and methods

### Studydesign

This is a Retrospective case –control study.

### Sample collection

A total of 102 samples were used in this study(66 cervical cancer tissue specimens and 36 extracted DNA from healthy controls,). The sample size was calculated using OSSE, an Online Sample Size Estimator. 68 only have been genotyped 42 cases and 27 controls. All specimens were obtained from the Department of molecular biology, institute of endemic diseases. The samples were prepared extracted DNA.

### Inclusion criteria

All samples that were diagnosed as cervical cancer and the positive detection of high risk (16,18) HPV by PCR.

### DNA quality and quantity

DNA concentration and purity were assessed by Nano-drop instrument. Then a concentration of 300ug/ul for working DNA was prepared from the stock DNA. The stock of DNA was stored at -20 C until used.

### Selection of TLR2 polymorphisms

TLR2 polymorphisms were selected from a literature review and the public database of TLR SNPs (Innate Immunity Program in Genomic Applications database; available at http://innateimmunity.net/). (18).

### Genotyping for TLR 2 (−196 to -174 del)

Allele specific Polymerase Chain Reaction (PCR) was carried out for detection of TLR 2 (−196 to -174 del) usingprimer sequences used for TLR2 (−196 to -174 del) were (F) 5’-CACGGAGGCAGCGAGAAA-3’ and(R)5’CTGGGCCGTGCAAAGAAG-3’ (19).PCR reaction was performed in total volumes of 25 μl. Master mix containing 7 μl of genomic DNA templates for 2μl of 10 mMdNTPs mix (2.5mM dATP, 2.5 mMdGTP, 2.5mM dCTP and 2.5 mMdTTP) 1.5 μl of 50 mM MgCl2, 2.5 μ l of 10 X PCR buffer (10 m MTris-HCl(PH 8.3) 50 mMKCl), 0.6 μ l of Taq polymerase (5 U/ul) 1 μ l forward primer and 1 μ l reverse primer. Volume was completed to 25 μl per reaction mixed with ddH2O. Negative controls for reagent contamination, consisting of all PCR reagents except template DNA, were run each time a PCR assay was performed with a known positive control for EB virus.

### PCR condition

PCR conditions is a modification of that described by Saumya et al(19). After initial 5-minute incubation at 95°C, the samples were subjected to 35 cycles of 30s minute at 95 C, 40s minute at 58 °C, and 40s minutes at 72°C. After the last cycle, the samples were held at 72°C for 7 minutes, and then cooled to 8 C.

### 3%Agarose gel preparation

1.5 grams of agarose was added to the mixture of 5ml of 10X TBE (0.023 M. Tris – Borate, 0.5 mMEDTA) buffer and 45 ml of distilled water (DW) in a flask, the mixture was melted in microwave oven for 2-5 minutes, after cooling gel down to 60-70 °C, 4 µl of ethidium bromide(0.5mg/ml) was added, then the 50 ml of molten cooled agarose was loaded without making air bubbles into the 50ml gel cast that has fixed spacers and combs and was left for 30 minutes for polymerization.

### Gel Electrophoresis

Running buffer composed of 5ml of 10X TBE buffer and 95ml of distilled water was powered in a gel cast. 5μL PCR product of each sample and the controls (Positive and negative) was mixed with 3μL loading dye (Bromophenol one in six)) on a sheet of parafilm, then was loaded into a well of agarose gel starting from lane 2 and 10μL DNA molecular weight marker 100 bp was loaded into lane 1. Lastly, tank was plugged into power supply and ran at 40 volts for 2 hours until loading dye has moved 3/4 the length of the gel. The gel was illuminated with UV transilluminator. a Gel-Doc System was used to store the image for further analysis.

### 8% polyacrylamide

After the PCR products were analyzed by 3%agarose the PCR product then analyzed by 8% polyacrylamide gel electrophoresis. a 286 bp band correspond del to TLR2 ins/ins, 264 bp to homozygous del while 286bp and 264 bp bands belonged to heterozygous individuals. The gel solution Prepared as follow:

30% Acrylamide (29:1)3.2 ml, H2O 6.4 ml,5x TBE 2.4 ml,10% APS 200 µl,TEMED 10 µl. quickly after addition of TEMED the gel completed before the acrylamide polymerized. then the appropriate comb inserted into the gel,. Then the acrylamide allowed polymerizing for 30-60 minutes at room temperature. After polymerization is complete, the gels inserted into Hoefergelbox. Then the running buffer was added and carefully the combs pulled from the polymerized gel the wells flushed out with 1x TBE. Then the DNA samples 10 μ l with the 2 μ l of gel loading buffer Loaded into the well. The electrodes Connect to a power pack, the power turned on 128V for 30S.

The gel immersed in 1XTBE (50ml) containing EtBr (0.5 µ g/ml) for 30 minutes at room temperature with gentleagitation. then the gel soaked in fresh DDI water for 15 minutes.

### Data analysis

Deviation from the Hardy-Weinberg equilibrium was tested using a chi square test. All statistical analyses were performed using Statistical Package for Social Sciences version 15.0. fisher exact test for the analysis of significant association. Statistical significance were two sided and taken as significant when p-value was less than 0.05.

## Results

### Age distribution of cases and control

In both cases and controls the age ranged from 21-85 with mean age of 60.5 in the cases and 50.5 in the control as shown in Figure 3-1, and Figure 3-2.

**Figure 3-1:**
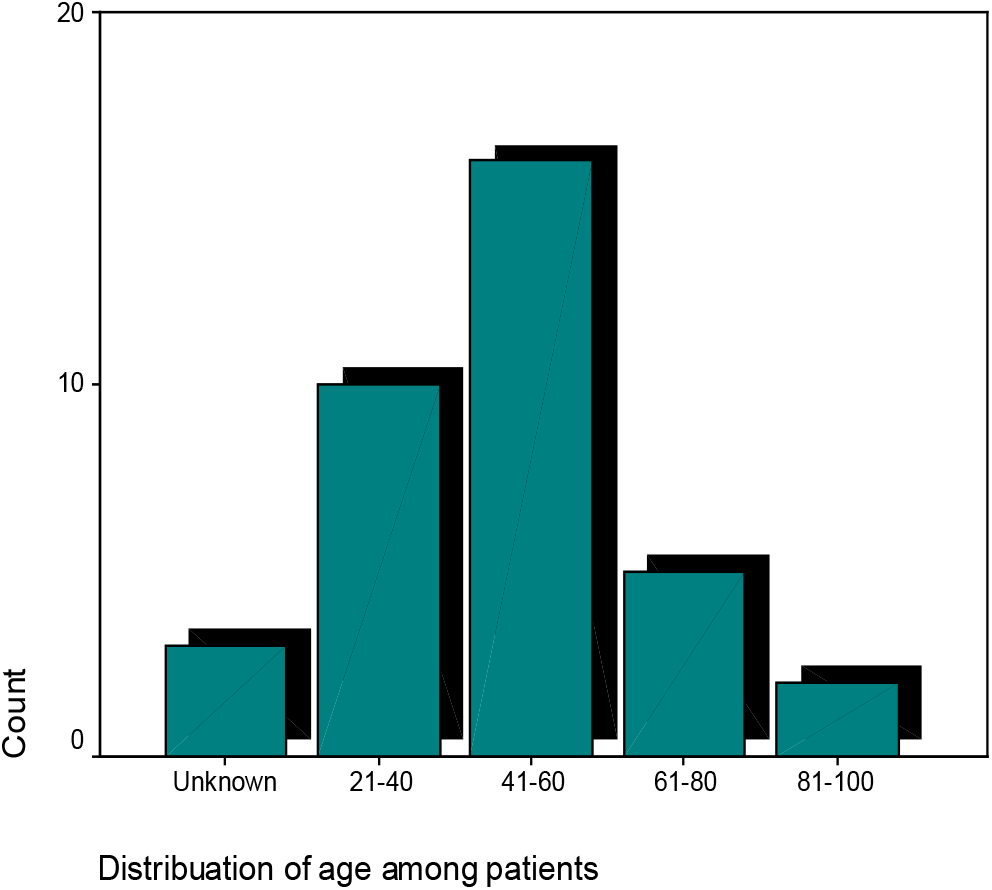
The distribution of age in cases

**Figure 3-2:**
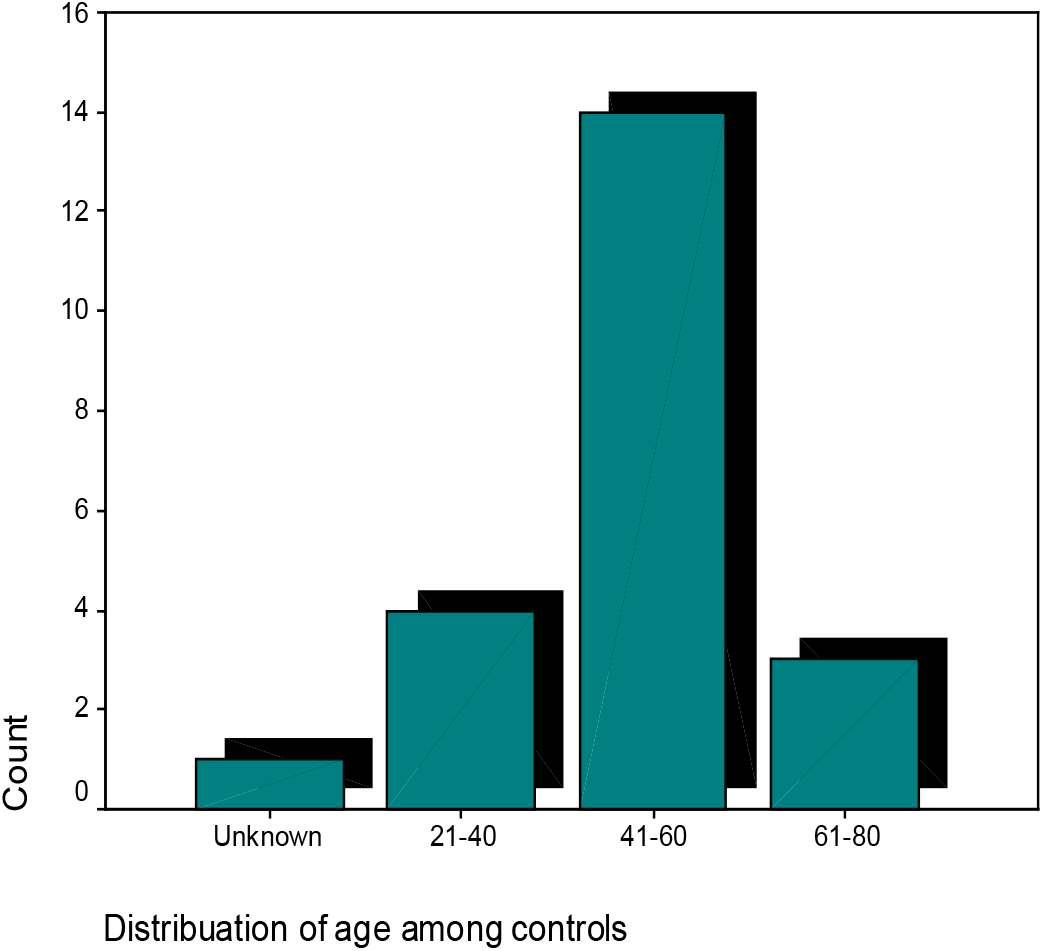
The distribution of age in control

### Genotyping of TLR2 polymorphisms

The genotype and allele frequencies of TLR2 gene, -196 to -173 del polymorphisms, were tested for HWE fitness. Cases& control were observed to be in Hardy– Weinberg equilibrium, (P>0.05in cases p-value = 0.840, p-value = 0.688 in control).

The TLR2 -196 to -173 wt/wt homozygote showed a single band of 287 bp, whereas the TLR2 -196 to -173 del/ del homozygote displayed a single band of 264 bp. The TLR2 - 196 to -173 wt/del heterozygote resulted in two bands of 287 and 264 bp are shown in (Figure 3-5).

**(Figure 3-3):**
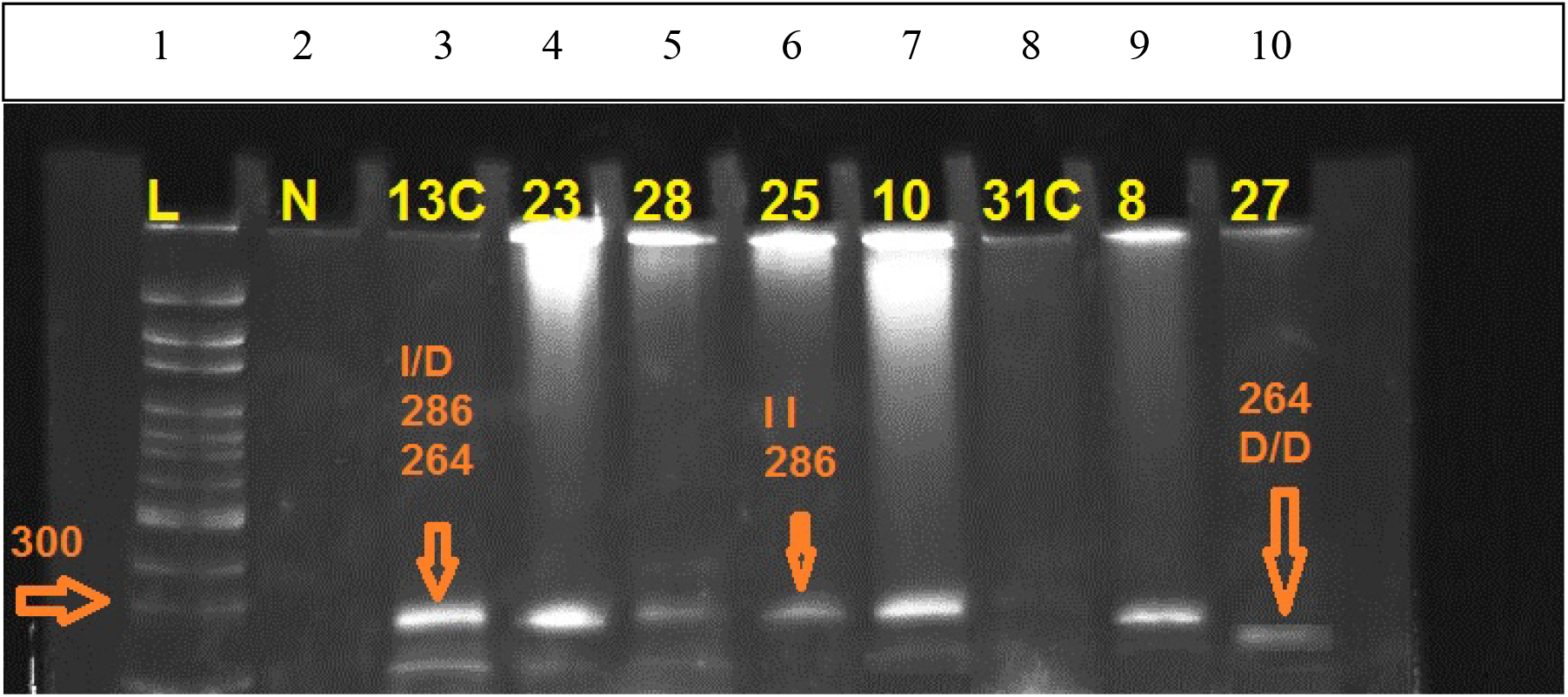
8% Polyacrylamide gel. Lane1 ladder(100)1300, Lane 2 negative control, Lane 3 control sample with genotype I/D, Lane 4,5,7,1,9 patient sample with genotype I/D, Lane 6 patient sample with genotype I/I, Lane 10 control sample with miner allele genotype D/D.

**Figure 3-4:**
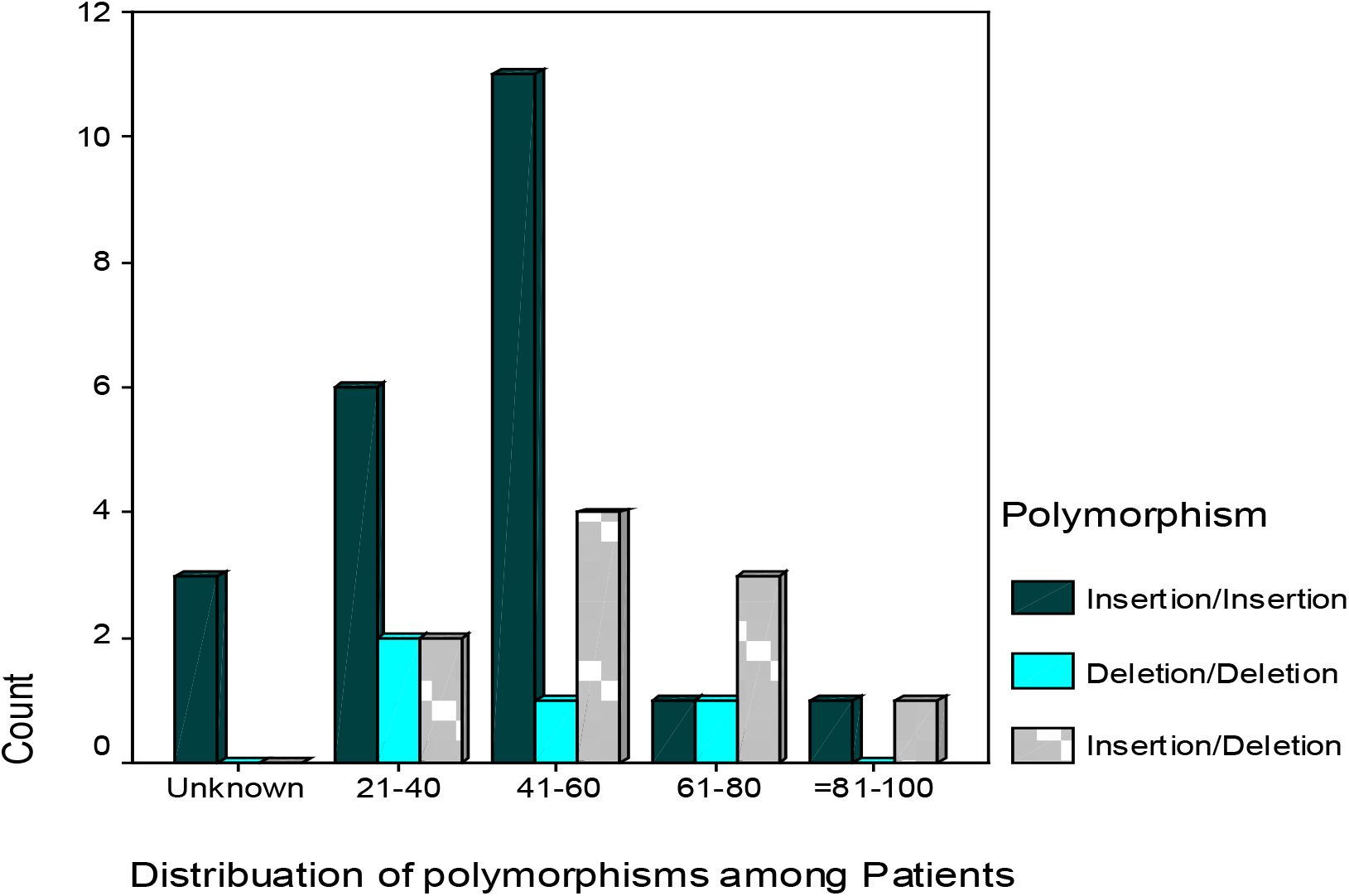
distribution of polymorphism in cases

**Figure 3-5:**
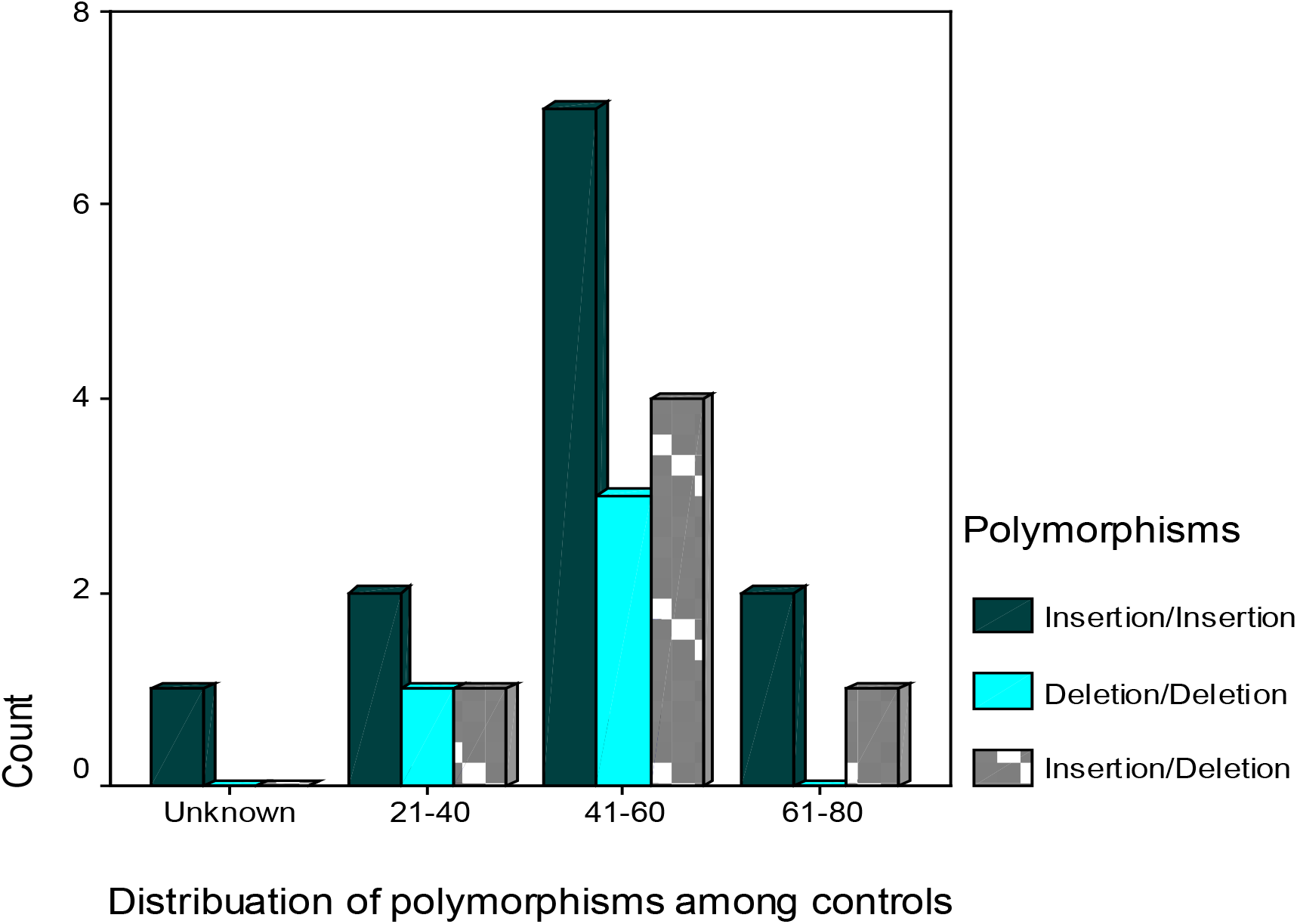
Distribution of polymorphism in control

### Polymorphism distribution among age groups in cases and control

Distribution of the TLR2 polymorphism among age groups is shown in (Figure 3-3) and, (Figure 3-4). The major age group in the control and cases was 41-60 years.

### Allele and genotype frequency

The allele frequency has been estimated for both cases and control as shown in (Table 3-3).

**Table 3-3:**
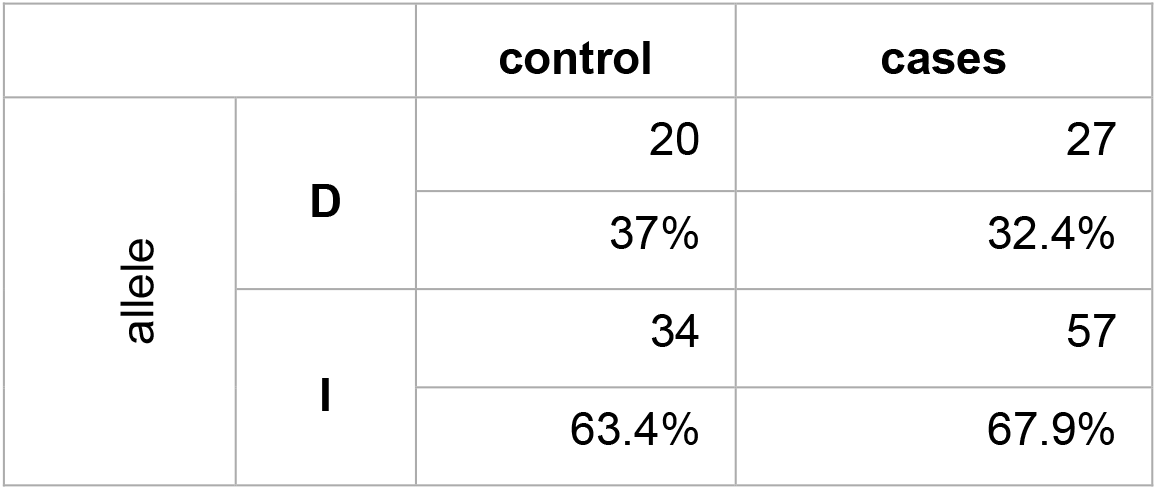
Allele frequency in cases & control

#### TLR2 genotype

The –196 to –174del polymorphism of *TLR2* was investigated in all 69 subjects. 27 were control subject 42 were cases. The distribution of *TLR2* genotypes in cases was ins/ins 47%, ins/del 38% and del/del 14%. The frequency of the del/del genotype seemed to be comparatively higher in our healthy control subjects25.9%, while the frequency of the haplotypewas considerably higher in cervical cancer cases.

No significant differences in –196 to –174 ins/del genotype (cervical cancer cases vs. healthy controls), OR = 0.536, 95% CI = (0.168 – 1.71) by Fisher’s exact test. similarly, no significance difference of –196 to –174 del /del genotype between cases and control.

### Annotation and functional assessment of genetic variants

Three-dimensional (3D) chromatin looping data http://cbportal.org/3dsnp/ were used to link promising SNPs to their three-dimensional interacting genes. As predicted by 3DSNP,rs111200466 is located in 28 transcription factor binding sites, it alters 2 sequence motifs, It locates in Enhancer state in 33 cell types.

## Discussion

Host genetic factors are emerging as key determinants of disease risk for many cancers. Identifying candidate genes is a significant challenge that has to stem from a profound understanding of the disease’s path physiology. Cervical cancer, a major health problem among women worldwide, is linked to persistent infection of HPV. It is believed that the virus’s presence is insufficient to induce carcinogenesis in the absence of other genetic changes within the cell (18).

Epidemiologic evidence reveals a putative link between inflammation and cervical carcinogenesis initiated by HPV infection (19); it has been proposed that inflammation acts as a cofactor for HPV (20).

Pattern recognition is an essential aspect of innate immunity. Cervical keratinocytes express Toll-like receptors (TLRs), which recognize (PAMPs) and function in immune surveillance in the cervix.. TLR2 is the most type of TLR expressed by fallopian tubes and cervix cells. Many SNPs have been identified in TLR2.

The -196 to -174 del is the most known SNP that affects its activity.

In this study, the TLR2 gene genotyped in 69 samples out of 102. The sample size of patients and control in the current study is possibly considered small for the analysis of polymorphisms. One Possible factor that limited this study was the quality of DNA of the samples. The DNA was extracted from paraffin block.

This study document that (TLR2 -196 to -174 del) is present in Sudanese women with allele frequency 37.0 in control (N=27) and 32.1 in cases (N=42). The genotype frequency of the D/D genotype was 14.3 in cases and 25.9 in control. The allele frequency was higher in control than the cases as well as the genotype frequency. However, no significant association was detected (p-value 0.520 OR=1.66) by fisher exact test and regression analysis.

Our result disagreed with the finding of a study done by Pandey S, where the del allele frequency was higher in cases with a significant association with the risk of cervical cancer in Indian women (17.7%in cases N=150 vs. 12.3% in control N=150, p=0.048) (21).In contrast to the finding of Pandey S, TomomitsuTahara, investigated the role of TLR polymorphism in susceptibility of developing gastric cancer after H.Pylori infection; he mentioned that D/D genotype is more frequent in control vs. cases.

Furthermore, the haplotype I/D genotype of Sudanese women demonstrates no difference in cases and controls (Table 3-4) ((38.1), (22.2) l), which was also reported by Pandey S and her colleagues, with I/D genotype frequency in cases and controls (28.7), (23.3) (21) respectively.

**(Table 3-4):**
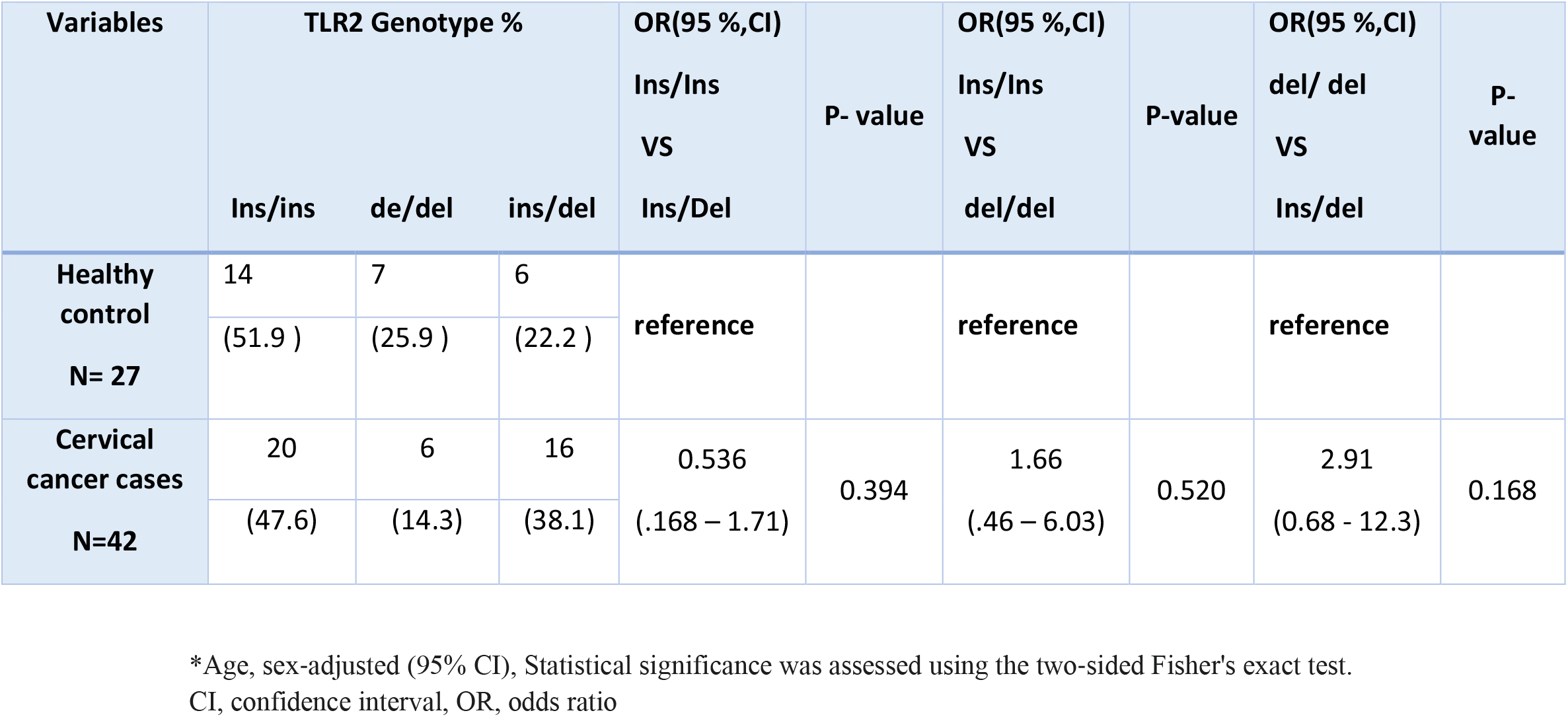
TLR2 polymorphism and risk of cervical cancer (compared with healthy control subjects)

| Variables | TLR2 Genotype % |  |  | OR(95 %,CI) | P- value | OR(95 %,CI) | P-value | OR(95 %,CI) | p- value |
| --- | --- | --- | --- | --- | --- | --- | --- | --- | --- |
|  | Ins/ins | de/del | ins/del | Ins/Ins<br>VS<br>Ins/Del |  | Ins/Ins<br>VS<br>del/del |  | del/ del<br>VS<br>Ins/del |  |
| **Healthy control**<br>**N= 27** | 14 | 7 | 6 | **reference** |  | **reference** |  | **reference** |  |
|  | (51.9 ) | (25.9 ) | (22.2 ) |  |  |  |  |  |  |
| **Cervical cancer cases**<br>**N=42** | 20 | 6 | 16 | 0.536 | 0.394 | 1.66 | 0.520 | 2.91 | 0.168 |
|  | (47.6) | (14.3) | (38.1) | (.168 – 1.71) |  | (.46 – 6.03) |  | (0.68 - 12.3) |  |

\*Age, sex-adjusted (95% CI), Statistical significance was assessed using the two-sided Fisher's exact test. CI, confidence interval, OR, odds ratio

As the D/D genotype has low activity and is more frequent in control, this may suggest a connection between TLR2 improper activation and protection from HPV persistency and cervical cancer. A comparison of cervical cytokine levels in women with persistent HPV infection and women with resolved HPV infections demonstrated more significant changes detected in the cervical mucus of persists than clearers. Cytokines that changed significantly throughout infection belong to the group of proinflammatory cytokines including IL-6, IL-2 increased significantly in persists but decreased in clearers at the first visit in which HPV DNA was detected (22). However, many studies indicated the high level of IL-10 in cervical cancer patients vs. control subjects, and IL10 is associated with disease progression (23). Many researchers have reported that TLR2 expression increased in cervical cancer samples compared to normal samples by the immunohistochemical method and real time PCR (24, 25).

Moreover, ArnabGhosha compared samples from individuals with normal, precancerous lesions, cervical intraepithelial neoplastic (CIN), and invasive squamous cell carcinoma (SCC) of the cervix. The expression of TLR2 correlated with disease progression, the noticeable and significant difference observed in late-stage invasive cases. Considering this finding and the fact that over activation of TLR may result in autoimmune, chronic inflammation, and neoplastic diseases. Therefore, Following TLR activation and the establishment of the inflammatory response against invading pathogens, there must be a checkpoint where TLR signaling is abolished. The system is returned to a normal physiological state to avoid a harmful response towards the host immune system. To date, numerous negative regulatory molecules have been characterized. One of these regulatory systems is transcriptional regulation (26). Disease severity is enhanced by increase expression of TLR2, so the presence of rs111200466 in the promoter region may interrupt this preferable context of highly expressed TLR2. We can suggest that rs111200466 inhibits or disrupt the negative control of tlr2 expression and activation. In our in silico annotation of rs111200466, as confirmed by 3DSNPs, this SNP position is a binding site of 28 transcriptional factors. One of these is BHIH40. In a study of tuberculosis and TLR2, the author found that these cells can sense Mtb via TLR2 and respond to the stimulation by secreting IL-10. IL-10 concentrations were significantly higher in Bhlhe40-/-cultures than in wild type cultures after heat-killed Mtb stimulation of these cells, suggesting that the system was relevant for studying how Bhlhe40 regulates IL-10 secretion in myeloid cells (27).

The study findings are supported by (Renee .2010) that showed that HPV’s clearance differs significantly with ethnicity. In African American participants, over half (57.5%) of HPV infections persist for 18 months or longer, while only 31.6% of HPV infections persist in European American participants, which indicates that genetic basis is implicated in cervical cancer susceptibility.

**In conclusion**, the TLR2 -196 to -173 del polymorphisms present an interesting target for investigating the genetic susceptibility to cervical cancer. Additional association studies of TLR6, a co-receptor of TLR2, and other candidate genes involved in immune response will be required to determine their contribution to cervical cancer susceptibility.

Besides, these polymorphisms in the TLR2 gene may influence the immune response to other pathogens recognized by TLR2. They may have important implications for understanding the pathogenesis of a wide range of infections and cancer in Sudan. This study is the first report of TLR2 -196 to -173 del polymorphisms in Sudanese women.

### Recommendation

Studies in a large and ethnically diverse population will be needed to resolve the impact of the TLR2 polymorphism in the susceptibility of cervical cancer in Sudan.

## Data Availability

all data in the manuscript are available

## Notes

### Competing Interest Statement

The authors have declared no competing interest.

### Funding Statement

no external funding was received

### Author Declarations

ethical committee institute of endemic diseases university of Khartoum

